# Transcutaneous afferent patterned stimulation reduces essential tremor symptoms through modulation of neural activity in the ventral intermediate nucleus of the thalamus

**DOI:** 10.1101/2024.12.02.24317799

**Authors:** Cuong P. Luu, Jordan Ranum, Youngwon Youn, Jennifer L. Perrault, Bryan Krause, Matthew Banks, Laura Buyan-Dent, Kip A. Ludwig, Wendell B. Lake, Aaron J. Suminski

**Author notes:** Address for Correspondence: Aaron J. Suminski 1111 Highland Ave, Rm. 3555 Madison, WI 53705.

## Abstract

Essential tremor (ET), the most common movement disorder in adults, presents with involuntary shaking of the arms during postural hold and kinetic tasks linked to dysfunction in the cerebello-thalamo-cortical (CTC) network. Recently, transcutaneous afferent patterned stimulation (TAPS), applied through a wrist-worn device, has emerged as a non-invasive therapy for medication refractory ET. However, its mechanism remains unclear. We hypothesize that TAPS reduces tremor through modulation of the VIM thalamus in the CTC network. Employing refractory ET patients seeking VIM deep brain stimulation (DBS), we quantified clinical tremor improvement following TAPS treatment in a pre-operative setting, followed by intra-operative, microelectrode recording of the contralateral thalamus with concurrent TAPS treatment on and off. After one preoperative session, TAPS significantly reduces upper limb tremor, with asymmetric effect favoring the treated limb and greatest improvement tending to kinetic tremor. The magnitude of TAPS-related tremor reduction demonstrates a positive correlation with the modulation of alpha and beta band LFPs in the VIM. TAPS also modulated spiking activity in the VIM, though it was uncorrelated with the degree of tremor reduction. Of note, TAPS related modulation of LFPs and spiking activity was greatest near the optimal placement location for DBS lead in treating ET. In sum, TAPS likely reduces tremor in ET by modulating the VIM and connected nodes in the cerebello-thalamo-cortical pathway.

## Manuscript

## Introduction

Essential tremor (ET) is the most common movement disorder in adults, and its impact is rising as a greater share of the world population age.^1,2^ ET patients experience debilitating symptoms from involuntary, rhythmic action tremor of the arms, though shaking can also involve the head, voice, and legs.^3,4^ These action tremors often involve two clinically distinct types with intrinsically different frequency: kinetic tremors, typically under 6 Hz, and postural tremors, which range from 6 to 12 Hz.^3,5–9^ Their exact etiology remains unclear, but, complementary evidence from postmortem histology^10,11^, imaging^12^, and electrophysiological^13^ studies point to dysfunction in the cerebello-thalamo-cortical (CTC) network—a network that links the deep cerebellar nuclei to the motor cortex via the motor and sensory thalamus, including the ventral intermediate (VIM) nucleus. The disruption of this network hinders one’s ability to evaluate and refine goal directed movement.^14–20^

There remains a significant gap in the treatment of ET. Frontline medical intervention with primidone and propranolol may improve tremor to a modest extent, but about 50% of patients stop medications due to side effects.^21,22^ Surgery for deep brain stimulation (DBS) targeting the VIM demonstrates a 60-75% reduction in tremor, yet up to 86% suffer ataxia, dysarthria, or dysgeusia and 15-73% eventually lose tremor control.^23,24^ The concerns of side effects, therapy habituation, and cost from DBS deter approximately half of eligible candidates.^25^ Thus, VIM DBS is generally reserved for patients with severe, debilitating ET. Recently, a less invasive alternative, magnetic resonance-guided focused ultrasound (MRgFUS), has shown promise in treating moderate to severe, refractory ET by creating a lesion in the VIM.^26–29^ Despite its safety and efficacy, MRgFUS adoption has also remain limited due to its need for specialized equipment, irreversible nature, and high relapse rate.^30^ Taken together, the limitations of current therapies leave many ET patients, especially those with mild to moderate symptoms, without effective treatment options.

In response to these limitations, transcutaneous afferent patterned stimulation (TAPS^TM^) emerged as a non-invasive, on-demand therapy to reduce hand tremor in ET patients.^31^ TAPS applies electrical bursts that alternate between the median and radial nerves on the wrist, leveraging afferent projections from these nerves to stimulate the VIM. Importantly, electrical stimulation of the median nerve is known to directly modulate neural activity throughout the somatosensory thalamus, including the VIM.^32–34^ The alternating burst of TAPS, whose frequency is tuned to the tremor frequency of each patient, is intended to disrupt synchronized, pathological firing of VIM neurons through a process known as coordinated reset.^35^ TAPS has gained FDA approval for ET treatment, demonstrating efficacy in a randomized controlled trial and a 38-83% tremor reduction lasting about one hour after each session.^36,37^ Its tremor-reducing effects have been shown to accumulate over months of treatment, with no habituation and minimal side effects.^38^

As proposed in theory, TAPS achieves tremor reduction by desynchronizing pathological, oscillatory neural activity in the CTC network.^39^ However, outside of data showing clinical improvement and a small FDG-PET/ CT study, there is limited evidence elucidating the underlying mechanism of TAPS and its potential impact on the brain.^40,41^ Here, we examined what effect TAPS has on the clinical components of ET and how it may modulate the CTC network. We hypothesized that TAPS would modulate the VIM thalamus in patients who see tremor reduction benefits. Our results provide strong evidence suggesting that tremor improvement resulting from TAPS therapy is related to modulation of neural activity in the CTC network, in particular, modulation of alpha band local field potentials (LFP).

## Results

### Study demographics, design, and TAPS

To prospectively study the effect of TAPS across tremor types and on thalamus activity, we enrolled 9 adults (2 female, Mean age ± SD: 65 ± 7 years) with medication refractory ET seeking DBS placement in the VIM. Demographic, medical history, and study-related measures are reported in Table 1. Using the Essential Tremor Rating Assessment Scale (TETRAS), all subjects presented with moderate hand tremor scores (Mean ± SD of spiral drawing score: 2.8 ± 0.8; upper limb tremor: 2.3 ± 0.5) approximating an average tremor amplitude of 5 cm. When surveyed with the Quality of Life in Essential (QUEST) questionnaire, tremor was present during most of patients’ waking hours (Mean ± SD: 82.1 ± 15.4%), with the greatest impact on hobbies/ leisure activities (Mean ± SD: 85.2 ± 15.5%) and general physical activities (Mean ± SD: 66.1 ± 25.2%).

**Table 1.** Demographic, Baseline Severity, and TAPS Settings.

| Subject | 1 | 2 | 3 | 4 | 5 | 6 | 7 | 8 | 9 | Cohort Summary |
| --- | --- | --- | --- | --- | --- | --- | --- | --- | --- | --- |
| Age (decade) | 50s | 70s | 70s | 60s | 50s | 70s | 60s | 60s | 60s | **65 ± 7** |
| Sex | M | M | F | M | M | M | M | F | M | **Male: 78%** |
| Age of Onset (decade) | 10s | 10s | 60s | 30s | 20s | 10s | 20s | 40s | 50s | **29 ± 17** |
| Family History | 1 | 1 | 0 | 1 | 1 | 1 | 0 | 0 | 0 | **56%** |
| **Specific Current Co-Therapy** |  |  |  |  |  |  |  |  |  |  |
| Propranolol | 1 | 1 | 1 | 0 | 0 | 1 | 0 | 1 | 1 | **67%** |
| Primidone | 1 | 1 | 1 | 0 | 0 | 0 | 0 | 0 | 0 | **33%** |
| Gabapentin | 0 | 1 | 1 | 0 | 0 | 0 | 0 | 0 | 0 | **22%** |
| **Initial QUEST Life Disability Scores**<br>**(Frequency Range: 0-100%)** |  |  |  |  |  |  |  |  |  |  |
| Tremor During Waking Hours | 89 | 83 | 56 | 89 | 89 | 89 | 94 | 94 | 56 | **82.1 ± 15.4** |
| Communication | 17 | 0 | 0 | 67 | 0 | 42 | 25 | 0 | 25 | **19.5 ± 23.2** |
| Work and Finance | 50 | 0 | 38 | 8 | 42 | 67 | 33 | 0 | 8 | **27.3 ± 24** |
| Hobbies and Leisure | 83 | 92 | 100 | 100 | 75 | 100 | 58 | 92 | 67 | **85.2 ± 15.5** |
| Physical | 64 | 28 | 100 | 64 | 31 | 97 | 75 | 78 | 58 | **66.1 ± 25.2** |
| Psychosocial | 61 | 17 | 58 | 33 | 39 | 31 | 64 | 28 | 8 | **37.7 ± 19.8** |
| **Initial TETRAS Scores**<br>**(Averaged Across Tasks, Severity Range: 0-4)** |  |  |  |  |  |  |  |  |  |  |
| Archimedes Spiral (Task 6) | 3.0 | 3.0 | 4.0 | 2.0 | 2.0 | 2.0 | 3.0 | 4.0 | 2.0 | **2.8 ± 0.8** |
| Upper Limb Tremor (Tasks 4, 6, 8) | 2.0 | 2.4 | 3.1 | 1.7 | 2.0 | 2.1 | 2.7 | 3.0 | 1.8 | **2.3 ± 0.5** |
| Total Task Score (Tasks 1-9) | 1.7 | 1.5 | 2.2 | 1.5 | 1.5 | 1.3 | 1.7 | 2.2 | 1.0 | **1.6 ± 0.4** |
| **TAPS Settings** |  |  |  |  |  |  |  |  |  |  |
| Dominant Hand | Right | Left | Right | Left | Right | Right | Right | Right | Right | **Right: 78%** |
| Tremor Frequency (Hz) | 5.4 | 4.5 | 4.5 | 4.5 | 5.7 | 8.1 | 5.3 | 6.5 | 4.1 | **5.4 ± 1.3** |
| Stimulation Current (mA) | 7.0 | 7.0 | 6.5 | 7.3 | 5.8 | 6.5 | 7.8 | 6.0 | 9.5 | **7.0 ± 1.1** |

Range displayed as mean ± 1 standard deviation.

Patients underwent the study in 2 phases, in which they donned a wrist-worn stimulator and received standard TAPS treatment on their dominant limb (Fig. 1A, B; see Methods 1.2). TAPS treatment was personalized for each participant at their tremor frequency (Mean ± SD: 5.4 ± 1.3 Hz) and at the maximum tolerable stimulation amplitude (Mean ± SD: 7.0 ± 1.1 mA), calibrated according to the manufacturer’s instructions. These stimulation parameters were held constant between both the pre-operative and intra-operative phases. Phase 1, pre-operative, further involved the assessment of tremor severity using the TETRAS before and after TAPS treatment (Fig. 1C). Phase 2, intra-operative, occurred during the stereotactic operation to place DBS electrode in the VIM, a standard surgery to treat refractory ET (Fig. 1D). Of note, we removed subject 8 from subsequent analysis because the individual reported an atypical, long-term tremor suppression for one month after their TAPS treatment in phase 1.

**Figure 1.**
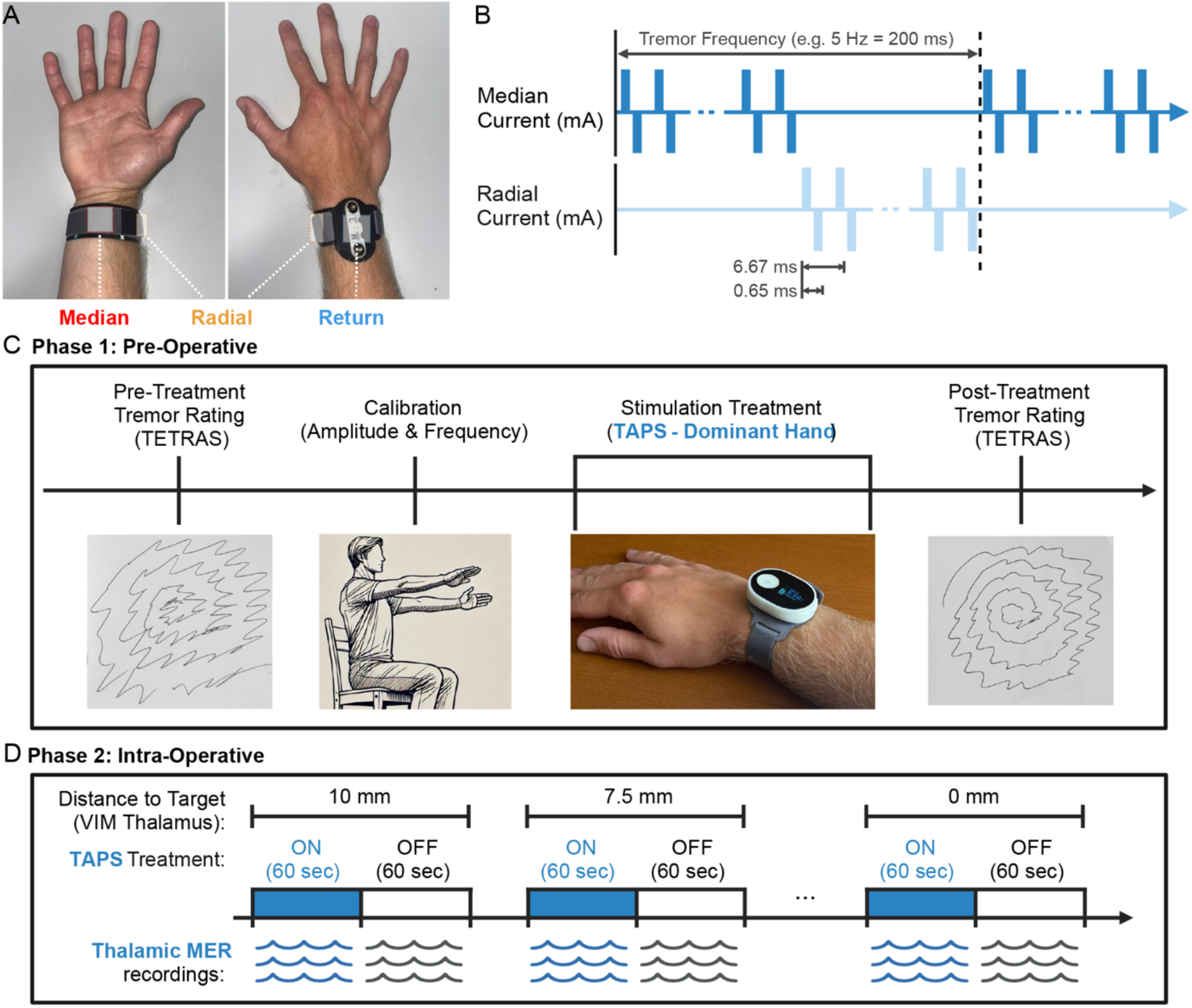
Investigating the effect of TAPS on clinical tremor scores and thalamus neural activity in essential tremor patients. **A** Demonstration of the wrist stimulation band placement on the dominant hand, showing the 3 electrodes used for the TAPS paradigm. Stimulator unlatched from the dorsal wrist, seen more clearly in **(C)**. **B** TAPS delivers alternating pulse trains to the median and radial nerves at a burst frequency matched to each patient’s tremor frequency. Stimulation amplitude is set to the maximum tolerable level reported by each patient. **C** During phase 1 (pre-operative), tremor severity is assessed using TETRAS scores before and after a 40-minute TAPS session. **D** During phase 2 (intra-operative), MER are collected along the planned DBS trajectory targeting the VIM nucleus of the thalamus, with TAPS alternating between the ON and OFF states.

### Individual and aggregated upper limb tremor tasks significantly improved in the dominant limb following TAPS treatment

Our first goal was to quantify the therapeutic effect of TAPS. We scored tremor severity using the 5 upper limb tasks in TETRAS before and after TAPS treatment, calculating tremor improvement as the difference in pre- and post-treatment scores (see Methods 1.3). Consistent with previous randomized controlled trials^36^, TAPS significantly improved the average total upper limb TETRAS score for the treated limb (0.61, *p*=0.008). Interestingly, we also found that TAPS resulted in a significant improvement in the average total upper limb TETRAS score of the untreated limb (0.35, *p*=0.008); however, improvement in the treated, dominant limb, was significantly greater than the untreated limb (*p*=0.047) (Fig. 2A). Given that ET is known to manifest with distinct postural and kinetic tremor components, we were also interested in examining TAPS related improvements in the individual upper limb tremor tasks. The treated hand showed a significant improvement in the forward postural hold (0.625, p=0.016), spiral drawing (0.875, p=0.039), and dot approximation (0.563, p=0.031) tasks with trends toward significant effects in the lateral postural (p=0.063) and kinetic (p=0.063) tremor measures (Supplementary Fig. S1). In contrast, the untreated limb showed no significant improvement in any of the individual tasks (p>0.05). We further analyzed handwriting, though separately as we did not collect handwriting data from the untreated, non-dominant limb, and found a significant improvement in handwriting tremor score resulting from TAPS (0.79, p=0.002) (Supplementary Fig. S2).

**Figure 2.**
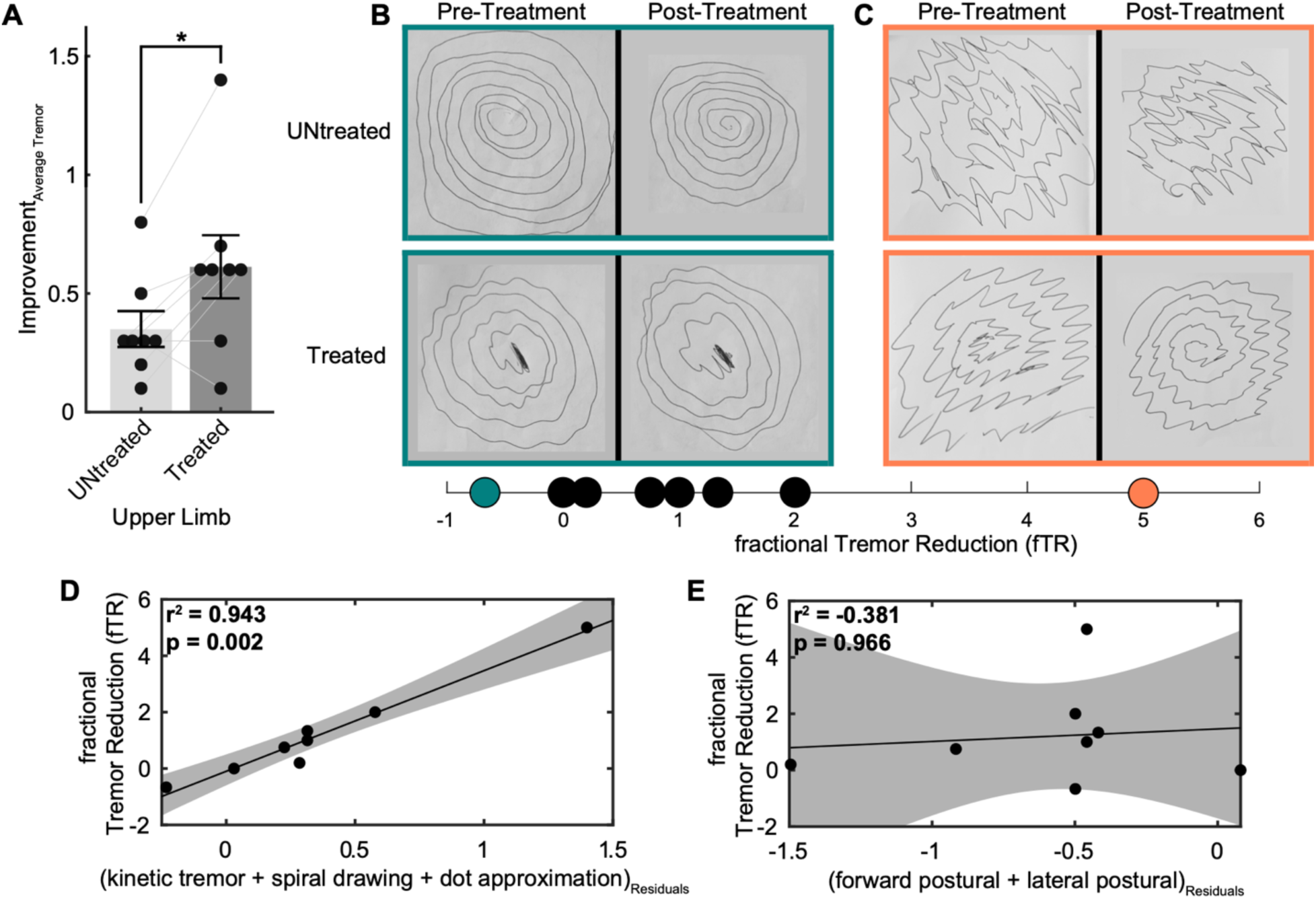
TAPS treatment improves essential tremor symptoms, with enhanced impact on kinetic tremor relative to postural tremor in the treated limb (n=8). **A** Mean ± SEM improvement in average upper limb tremor scores after 40 minutes of TAPS treatment, comparing the treated versus untreated limb across all patients (*two-sided signed-rank p=0.029). **B–C** Representative examples showing changes in spiral consistency and amplitude following TAPS treatment in patients with **(B)** low fTR (cyan) and **(C)** high fTR (orange). **D–E** Multiple linear regression analyses showing the relationship between fTR and aggregated improvement in **(D)** kinetic tremor tasks and **(E)** postural tremor tasks. To visualize the regression model, the x-axis rescales the combined effect of all independent variables, adjusted using the Frisch–Waugh–Lovell theorem (Matlab fitlm).

### TAPS therapy enhanced improvement of kinetic tremor in the dominant limb

Given the significant reduction in tremor severity observed for both limbs, we were interested in isolating the effect of TAPS to the treated limb. We captured this through the fractional tremor reduction (fTR) score, calculated as a difference in average upper limb tremor improvement between the treated and untreated limb with normalization to the untreated limb value (Eq. 1). fTR ranged from -0.67 to 5.00, showing that subjects’ responses to TAPS had a wide dynamic range corresponding to observed qualitative tremor improvement (Fig. 2B, C). Here, half of the subjects exhibited fTR≥1 (i.e. double the tremor improvement in the treated limb compared to the untreated limb).

To continue the examination of how TAPS may impact the two components of ET, we explored the differential impact of TAPS on tasks that were grouped into either the kinetic or postural tremor components. We questioned if initial tremor severity correlated with fTR as tremor severity may be predictive of ET onset and progressive atrophy along neural pathways.^42,43^ We used the pre-treatment TETRAS scores as a proxy for initial tremor severity. Regression analysis found no relationship between fTR scores and tremor severity for postural or kinetic tremor (Fig S3). Next, we explored if TAPS exhibited differential improvement in either kinetic tremor or postural tremor. We discovered that improvement in kinetic tremor scores of the treated limb displayed a significant positive relationship with fTR, explaining 94.3% of the variability (F(3,4)=39.623, p=0.002) (Fig. 2D). In contrast, there was no relationship between improvement in postural tremor and fTR (p=0.967) (Fig. 2E).

### Fractional tremor reduction in the dominant limb directly correlates with LFP modulation in the VIM

Next, to understand TAPS’s therapeutic mechanism, we investigated if therapeutic effect was mediated by changes in neural activity within a prominent node in the CTC network, the VIM. During each patient’s awake DBS implant surgery, we advanced 3 microelectrodes along a trajectory targeting the VIM, recording at selected depths in and around the VIM while TAPS treatment was ON and OFF. Subsequently, we processed the recordings for the power of the LFP at different frequency bands from 0 to 200 Hz, and quantified the modulation index of LFP (MI_LFP_) as the fractional difference in power between the ON and OFF states (Fig. 3A, B). Since we suspected that TAPS may act on the same neural substrate as DBS, we performed an initial survey of the relationship between MI_LFP_ and distance from the final implant location of the DBS lead (i.e. the distal edge of the most distal contact on the DBS lead; see Methods 1.4).

**Figure 3.**
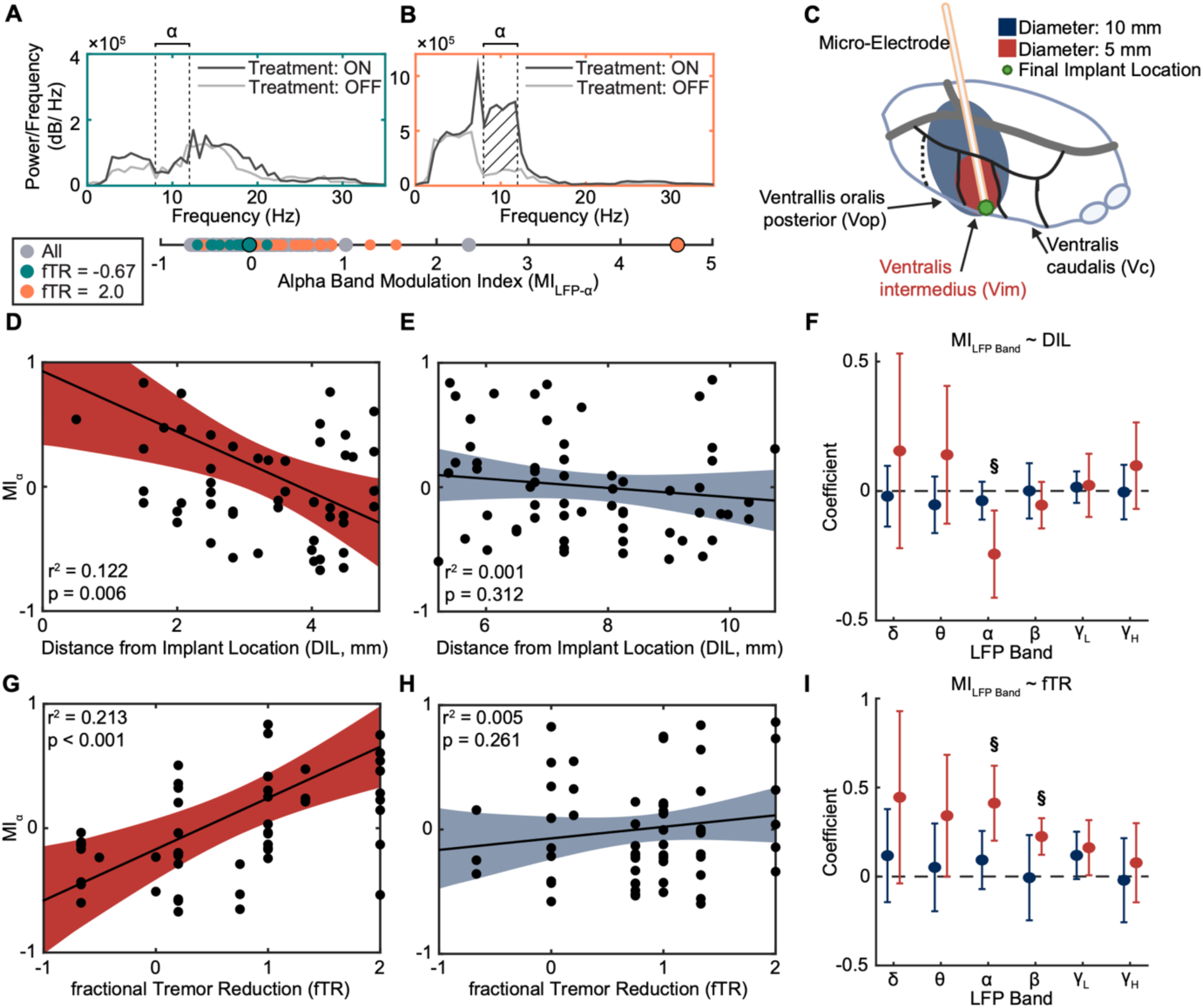
TAPS treatment modulates thalamic local field potentials with varied effects based on location and degree of tremor control. **A–B** Representative power spectral density (PSD) estimates comparing **(A)** low α-band modulation index (MI_LFP-α_) in a patient with low fTR (cyan) versus **(B)** high MI_LFP-α_ in a patient with high fTR (orange). Black and gray lines show Treatment ON and OFF conditions, respectively. **C** Schematic illustrating MER distance relative to the final implant location within the thalamus (defined as the distal edge of the DBS lead’s distal contact). MER locations are classified as within VIM (red) or outside VIM (blue). **D–E** Correlation between MI_LFP-α_ and distance from implant location (DIL) for recordings within VIM (red) and outside VIM (blue) (n=8). Points indicate MER data from individual depths, superimposed by best-fit lines and 95% confidence intervals. **F** Mean regression slope coefficients ± 2 SEM showing the relationship between MI_LFP-Band_ and DIL for recordings within VIM (red) and outside VIM (blue). ^§^ indicates regression slope coefficient with p≤0.008 (Bonferroni-corrected α’ threshold). (See supplementary Fig. S5 for expanded regression plots.) **G–H** Correlation between MI_LFP-α_ and fTR for recordings within VIM (red) and outside VIM (blue). **I** Mean regression slope coefficients ± 2 SEM showing the relationship between MI_LFP-Band_ and fTR for recordings within VIM (red) and outside VIM (blue). ^§^ indicates regression slope coefficient with p≤0.008 (Bonferroni-corrected α’ threshold). (See supplementary Fig. S6 for expanded regression plots.)

Regression analysis, however, found no relationship between MI_LFP_ in any frequency band and distance from the DBS lead implant location (Supplementary Fig. S4). We examined this relationship more deeply by parsing MER recordings that were putatively made within the VIM (Fig. 3C, ≤ 5mm from implant location) or outside of it (>5mm). Here, regression analysis found that, only within the VIM, the alpha band MI_LFP_ was strongly modulated closer to the DBS implant location (R2=0.1221, F(1,52)=8.371, p=0.006) (Fig. 3D, F). In contrast, no relationship between MI_LFP_ and distance to implant location was found outside of the VIM in any frequency band (Fig. 3 E, F; Supplementary Fig. S5), suggesting that TAPS and DBS may share a common mechanism for mitigating tremor.

Since tremor improvement due to TAPS, measured by fTR, varied widely among patients, we examined if there was a relationship between fTR and changes in neural activity quantified by MI_LFP_ across different frequency bands. Within the VIM, regression analysis found that patients who experienced the largest fTR due to TAPS had significantly increased MI_LFP_ in only the alpha (F(1,52) =15.313, p<0.001) and beta bands (R2=0.255, F(1,52)=19.16, p<0.001) (Fig. 3G, I). For recording locations outside the VIM, there was no relationship between tremor reduction and MI_LFP_ for any band (Fig. 3H, I; Supplementary Fig. S6). Notably, this effect was specific to tremor reduction as measured by fTR; a separate regression analysis showed no relationship between MI_LFP_ within the VIM and total tremor improvement in the treated upper limb (i.e. no normalization by effect in the untreated limb).

### TAPS suppresses multiunit spiking in the VIM, though effect was homogenous among patients

In addition to thalamic LFP, we also examined the modulation of multiunit spiking activity in response to TAPS treatment. One representative sample, recorded within the VIM 0.5 mm from the final implant location, showed a pronounced reduction in spike rate during TAPS treatment (Fig. 4A, B). We limited our analysis to recording depths within the VIM (i.e. ≤5 mm from the final implant location) where the minimum firing rates was at least 1 Hz while TAPS was OFF. We used a resampling procedure to estimate the most likely multiunit firing rate. This was necessary because many time series had different durations due to removal of motion artifacts (Fig. 4C, D). In the 37 recordings that met this criterion, we characterized the modulation index of spikes (MI_Spikes_) as the fractional difference in spike counts between the ON and OFF states of TAPS. MI_Spikes_ significantly decreased closer to the final implant location, with trend toward MI_Spikes_= -1 indicating that TAPS suppressed spiking activity to near 0 (R2=0.104, F(1,35)=5.197, p=0.029) (Fig. 4E). No significant relationship was observed between MI_Spikes_ and isolated tremor improvement (p=0.777) (Fig. 4F). At distances >5mm, MI_Spikes_ ranged from - 1 to 0.5, but it did not exhibit a significant relationship with either distance from implant location or isolated tremor improvement (p=0.768 and p=0.949, respectively).

**Figure 4.**
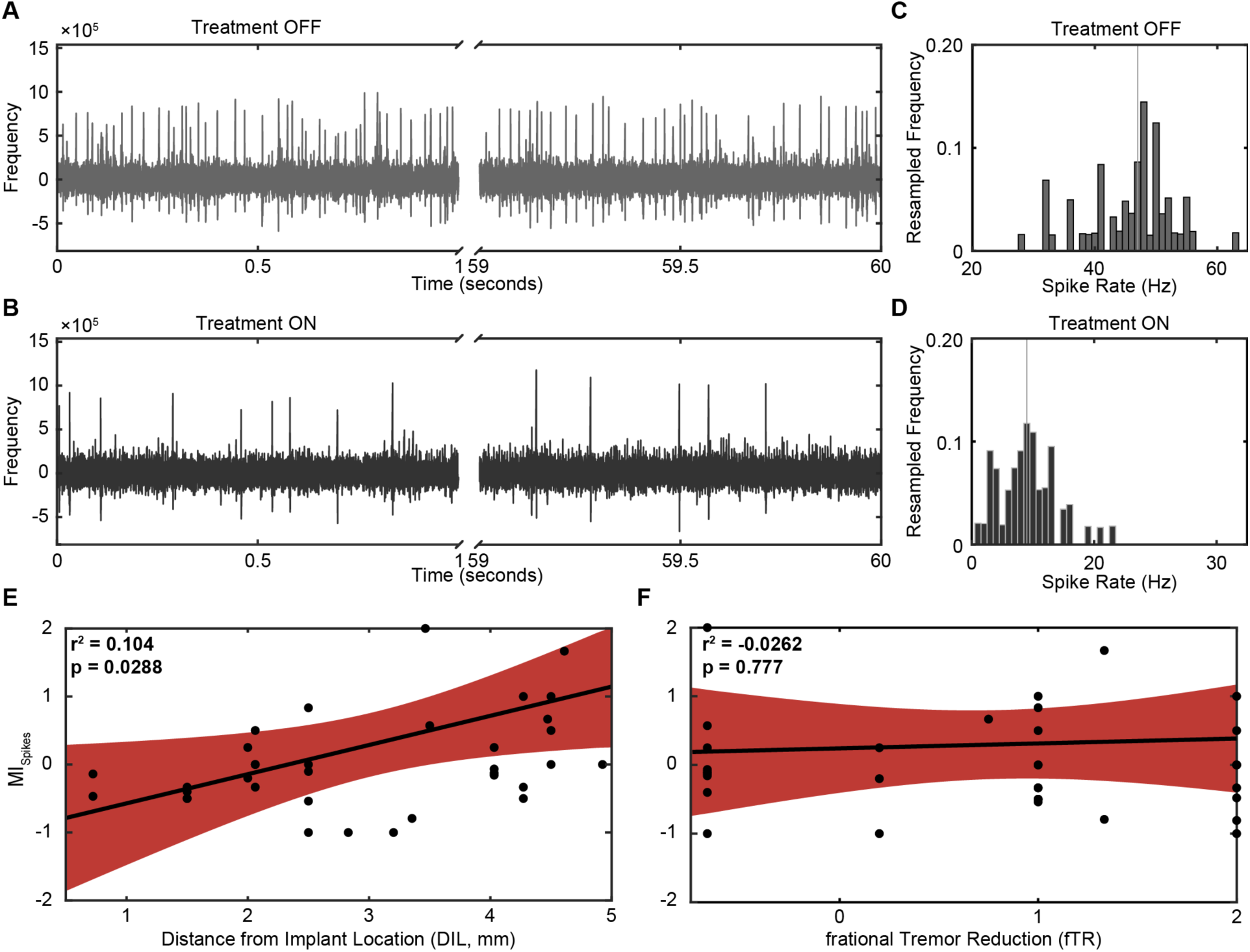
Modulation of spiking activity in thalamic MER under TAPS treatment ON and OFF conditions. **A–B** Sample time series of multiunit activity recorded 0.5 mm from the final implant location within the VIM thalamus during TAPS treatment **(A)** OFF and **(B)** ON periods. Spike rates (Hz) were calculated in consecutive, non-overlapping 1-second segments of the recordings, then aggregated for bootstrap resampling. **C–D** Spike rate distributions obtained through 10,000 bootstrap resampling iterations of the rates collected from recordings shown in **(A)** and **(B)**, during TAPS **(C)** OFF and **(D)** ON conditions, respectively. Vertical lines indicate median spike rates, representing the most likely spike rate for each recording. **E–F** Simple linear correlations between MI_Spikes_ and **(E)** DIL or **(F)** fTR. Data points represent individual recordings taken ≤5mm from the implant location (red) with baseline spike rates >1Hz during TAPS OFF. Best-fit lines with 95% confidence intervals are shown.

## Discussion

In the effort to find non-invasive therapy for ET, transcutaneous afferent patterned stimulation of the median and radial nerves near the wrist (i.e. TAPS) has been shown to reduce limb tremors, but its mechanism of action remains unclear. Putatively, stimulation alternating between the median and radial nerves serves to provide a desynchronizing input to the sensorimotor thalamus (i.e. VIM) via projections ascending in the dorsal column and/or spinothalamic pathways. The VIM links cerebellar output with the sensorimotor cortex and serves as a major node in the CTC network, the hypothesized locus of dysfunction in ET^10–13^. Given this anatomical substrate and that lesional therapies^26–29^ and DBS^23,24^ target the VIM for treatment of ET, we hypothesized that VIM is a likely site of the therapeutic effect of TAPS. Here, we demonstrated that TAPS of the median and radial nerves directly modulate the activity of both local field potentials and spiking activity in the VIM thalamus. Specifically, our data demonstrates that patients who have the greatest modulation of alpha and beta LFP caused by TAPS experienced the largest reduction in tremor symptoms. Additionally, sites exhibiting the largest modulation of both LFP and spiking activity are generally located closest to the placement of the DBS lead for treatment of ET (annotated as the distance from implant location and marked by the distal edge of the distal electrode on the DBS lead). Finally, our data further suggested that TAPS have an asymmetric effect on tremor reduction in different subtypes, with an enhanced effect on kinetic tremor relative to postural tremor in the treated limb.

### TAPS acts on the cerebello-thalamo-cortical network

There is strong evidence linking dysfunction throughout the CTC network, particularly thalamocortical circuits, to ET.^13,44–48^ For example, Kane et al. reported increased theta (4-8Hz) and alpha (8-12Hz) LFP power in thalamic nuclei receiving cerebellar input (i.e. VIM) compared to nuclei receiving input from the basal ganglia or somatosensory afferents. In conjunction, LFPs recorded from the VIM demonstrated increased synchrony in ET patients compared to individuals diagnosed with other neurodegenerative diseases^46^, and motor cortex alpha LFP power is altered in ET patients when compared to healthy controls.^13^ Moreover, electrical stimulation of the median nerve is known to modulate neural activity throughout the CTC network with the most direct effects being found in the somatosensory thalamus.^32–34^ Our data bridge the evidence describing CTC network dysfunction in ET and a putative therapeutic mechanism for TAPS, that is the modulation of neural activity in VIM. Notably, we show that TAPS of the median and radial nerves increased modulation of VIM alpha and beta bands LFP in subjects with higher fTR (Fig 3G, I), indicating a mechanistic role for VIM in reducing tremor. This effect was limited to the VIM as no LFP modulation was observed at distances >5mm from the location of the implanted DBS lead (Fig 3E, H). Notably, modulation of multiunit spiking activity was also only observed within the VIM; but, unlike alpha and beta LFP, had no relationship to reduction of tremor symptoms (Fig 4E, F). The dissociation between LFP and spiking activity modulation’s relationship to tremor reduction (i.e. fTR) is particularly interesting and potentially meaningful given their distinct roles.^49^ Specifically, Low-frequency field potentials typically reflect summed synaptic currents—potentially representing inputs to an area—while single/ multiunit activity likely represents outputs. In this light, the mechanism of TAPS may be thought to act on the CTC network more broadly via the VIM, and not on local neural populations in the VIM alone.

In validating our neural data, we believe that these results are not confounded by artifacts from TAPS applied at the wrist. First, raw time series showed no evidence of stimulation artifacts (Fig 4A, B; Supplementary Fig. S8). Second, modulation of neural activity related to tremor reduction (i.e. fTR) was only found in a spatially defined region, putatively within the VIM. Given the small range between the most dorsal and ventral recording locations (∼10mm), we would expect any presenting stimulation artifact to appear at all depths—that was not seen. Finally, the wrist stimulation device is wireless and battery-powered with a localized, bipolar electrode geometry, mitigating the effect of ground loop contamination and broad spread of the applied electric field.

### Heterogeneous tremor improvement in response to TAPS treatment

In our study, we observed that TAPS significantly improved tremor in both the treated *and* untreated limb. As described above, the effect of TAPS is putatively due to modulation of the CTC network via the VIM contralateral to the treated limb. The observed decrease in tremor in the untreated limb, however, is unexpected and its origin is unclear. Anecdotal observations and some reports have suggested that unilateral VIM DBS results in a significant tremor reduction in the arm ipsilateral to the implant^50^, but this ipsilateral benefit of DBS remain controversial and inconclusive.^51–53^ It is possible that the tremor improvement observed in the untreated limb of our patients may have resulted from a sham/ placebo effect. This interpretation is based on results from recent randomized controlled trial studying the effect of TAPS.^36^ Here, Pahwa et. al reported that subjects who had received TAPS treatment displayed tremor improvement similar in magnitude to what we see in the treated limb of our subjects (Fig. 2A). Of significance, we see further concordance in the magnitude of tremor improvement between Pahwa et. al’s subjects who received sham treatment and the untreated limb of our subjects. Thus, we believe that the improvement in the untreated limb, while statistically significant, is a sham effect. However, we did not test sham-treatments to provide conclusive evidence; future sham-controlled studies assessing tremor improvement in both the treated and untreated limb should be performed to demonstrate if unilateral TAPS exerts a bilateral effect on upper limb tremor.

Due to the unknown origin of tremor reduction in the untreated limb, we used a fractional difference measure, fTR, to estimate the effect of TAPS in the treated limb over and above the effects common to both limbs. Importantly, tremor improvement scores computed with fTR stratified patients into low and high responders in a similar proportion to previously published methods (i.e. half of the patients were high responders with fTR≥1).^37,38^ We found that tremor reduction measured by fTR was primarily driven by improvements in kinetic tremor (assessed by spiral drawing, finger-to-nose movement, and dot approximation) rather than postural tremor. This is an interesting result especially considering that kinetic and postural tremors have different characteristic frequencies of oscillation, <6Hz and 6-12Hz, respectively.^3,7–9^ It is possible that the alternating burst frequency of TAPS could modified to directly target various tremor subtypes and thereby personalize treatment based on an individual’s symptom profile.

Consistent with prior studies exploring tremor reduction due to TAPS^36–38^, we found that patients experienced varied degrees of tremor improvement. We explored a few possible explanations for the observed heterogeneity in response to TAPS. First, it is possible that engagement of the median and radial nerves at the wrist varied between patients; improper alignment of the stimulation electrode with the underlying anatomy or inadequate stimulation intensity could have diminished therapeutic effects. Here, wristband sizing, positioning, and stimulation parameters for TAPS were determined based on manufacturer’s instructions (i.e. tremor frequency measured during postural hold and stimulation amplitude set as the maximum tolerable current for 40 minutes of treatment). Importantly, all subjects verified the presence of paresthesia, not muscle contraction, in the median and radial nerves’ hand dermatomes with TAPS activation during both phases of the experiment. The presence of paresthesia referred to the fingertips, lacking reported pain or visible muscle recruitment, indicates engagement of large myelinated proprioceptive and cutaneous afferents (A-alpha and A-Beta fibers) in the median and radial nerve trunks.^54,55^ Therefore, subjects likely experienced adequate target nerves engagement. Second, dysfunction in the transmission of sensory information from the peripheral nerve to the central nervous system could contribute to varied efficacy of TAPS. Such dysfunction may be due to the severity of disease or another unknown factor. Importantly, our data showed no relationship between fTR and pre-treatment tremor severity, and no patients in our cohort reported sensory dysfunction. Thus, we expect sensory dysfunction did not impact our results. We did not, however, explicitly characterize proprioceptive or somatosensory acuity in our patients, and are not able to definitively rule out sensory deficits as a factor in the heterogeneous response to TAPS. Thus, relationship between the sensory perception of TAPS and the degree of tremor reduction remains an open question. Full exploration of the effect of the perceived sensation and its relationship to fiber type recruitment will likely require further extra-operative experiments.

### Limitations

We acknowledge that there are several limitations regarding this work and the conclusions drawn. First, our small cohort size reflects the exploratory nature of this research, as TAPS’s effects on CTC network activity had not been previously documented. Future confirmatory studies with pre-registered methodology will be essential to validate and expand on the findings elaborated here. Second, research using intraoperative neural recordings faces inherent limitations due to the clinical priorities of the surgery. For example, the use of sedatives during the DBS procedure may have influenced microelectrode recordings. This concern is likely muted, however, due to the rapid clearance of remifentanil and its minimal effect on neural activity.^56,57^ In addition, we also limited surgery duration to minimize infection risks and patient discomfort; thus, we could not test multiple stimulation parameters at each recording depth. Instead, we focused on recording neural activity resulting from TAPS treatment at a single current amplitude and alternating burst frequency. Future work that aims to explore or optimize the effect of stimulation parameters on VIM neural activity should employ chronically implanted recording methods. Such methodology would track long-term changes, enabling more robust analyses to address the cross-sectional limitations of this study. For example, larger datasets from such studies could also support advanced machine learning models to predict and optimize TAPS-related tremor reduction on an individual basis. Third, the lab-based quantification of TETRAS tremor scores may have induced stress in patients through a “white coat effect,” potentially biasing our measures of TAPS efficacy compared to home use and contributing to the observed response heterogeneity. Though, this is unlikely because the heterogeneity in tremor reduction captured by fTR matches the spread in therapeutic effect seen by patients with long-terms home use of TAPS.^38^ Finally, our results should be applied with caution when placed in the context of other non-invasive therapies using peripheral nerve stimulation to treat the symptoms of ET as the mechanism of action described here may be unique to TAPS.

## Conclusion

TAPS significantly reduced upper limb tremor severity in our cohort of medically refractory ET patients, with the treated limb showing greater improvement in kinetic tremor tasks. The therapeutic effect was associated with modulation of alpha and beta band local field potentials in VIM thalamus, with the greatest modulation proximal to the DBS implant site. In contrast, while VIM multiunit spiking activity was also modulated, these changes did not correlate with tremor reduction. This dissociation between field potentials and spiking activity suggests TAPS may achieve therapeutic benefit by modulating multiple nodes of the CTC network connected to VIM, potentially including the motor cortex and cerebellum, rather than through direct effects on VIM alone.

## Material and methods

### 1.1 Subject Inclusion & Exclusion Criteria

We prospectively enrolled 9 adults with medication refractory ET seeking DBS placement in the VIM. Key inclusion criteria were (1) age between 18 and 85 years old, and (2) a baseline, dominant hand score of 2+ on TETRAS item 6 Archimedes spiral drawing. Spiral drawing score served as one proxy for initial tremor severity, where a score of 2+ was selected to match the severity of subjects recruited in past studies.^36,58^ Subjects were excluded if they (1) were pregnant or (2) had cognitive disability impairing understanding of the consent process or experimental directions. All subjects provided written informed consent. The study was approved by the Institutional Review Board (IRB) at the University of Wisconsin – Madison (IRB ID: 2018-1052).

### 1.2 Transcutaneous Afferent Patterned Stimulation (TAPS)

We fitted each subject with a wrist-worn stimulator (Cala TWO band, Cala Health, CA, USA) to deliver TAPS on their dominant upper limb. The band hosted pad electrodes targeting the median and radial nerves, in addition to an electrode on the dorsal wrist for electrical return (Fig. 1A). After donning the device, the maximum tolerable stimulation current amplitude (mA) and alternating burst frequency (Hz) were calibrated for each subject. To attain the maximum tolerable current, stimulation was increased in 0.25 mA increments until the subject reported paresthesia in both the median and radial nerves’ hand dermatomes. The final current was the highest amplitude the subject reported that they could tolerate for 40 minutes. As for the alternating burst frequency, it was measured by on-board accelerometry and determined as the higher of two tremor frequencies after each subject had performed the forward and lateral postural hold tasks. The alternating burst frequency controlled how many alternating stimulation sets are delivered to the median and radial nerves per second (Fig. 1B). Stimulation on each of the median or radial nerve was delivered in biphasic pulses at 150 Hz, each phase lasting 300 µs with a 50 µs interphase period.

### 1.3 Phase 1: Pre-operative Quantification of Tremor Changes Following TAPS

Tremor severity was evaluated using TETRAS, picked for its high sensitivity to tremor changes and reliability between raters.^7^ TETRAS contained 9 performance items to rate action tremor in the head, face, voice, limbs and trunk from 0 (no tremor) to 4 (severe tremor) in increments of 0.5. Tremor exhibiting a higher amplitude (cm) scored higher in severity. In addition, tremor impact on life quality were also surveyed using the QUEST as these measures hold a modest insight into tremor severity.^59^ We scored tremor severity for the dominant and non-dominant limb using TETRAS before and after 40 minutes of TAPS delivered to the wrist of the dominant limb (Fig. 1C). Total upper limb tremor improvement was taken as an average of the differences in scores before and after treatment in 5 tremor tasks described by TETRAS items 4, 6, and 8 (Eq. S1). TETRAS item 4 tested (1) forward postural, (2) lateral postural, and (3) kinetic finger-to-nose tremors; item 6 tested (4) spiral drawing; and, item 8 tested (5) dot approximation tremor.

#### 1.3.2 Characterizing Tremor Improvement in the Treated Limb

In addition to computing tremor improvement for each limb separately, we were interested isolating the effect of TAPS in the treated limb from those that were potentially common to both limbs. Here, we used tremor improvement in the untreated limb as a within-subject normalization factor for the quantification of how TAPS affected tremor in the treated limb. Subsequently, the fTR of the treated limb following TAPS was a computed as a fractional change in tremor improvement of the treated limb relative to the untreated limb:

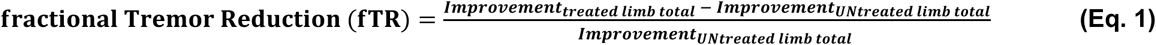

Therefore, fTR estimates the effect of TAPS in the treated limb over and above the effects common to both limbs.

#### 1.3.3 Comparative Analysis of Clinical Tremor Improvement

Improvement (i.e. the difference between pre- and post-treatment TETRAS score) of the 5 individual tremor tasks and the average total score were compared to zero and between limbs using two-sided Wilcoxon signed-rank tests (α=0.05). To explore the differential impact of TAPS on kinetic and postural tremors and how the improvement of each contributed to overall fTR, measures of kinetic finger-to-nose tremor, spiral drawing tremor, and dot approximation tremor were classified as kinetic tremor while forward postural and lateral postural tremor were grouped as postural tremor.^6^ Subsequently, a multilinear regression (Matlab fitlm function) was used to assess the relationship between fTR and the improvement in either kinetic or postural tremors (Supplementary Eq. S2 A,B, α=0.05). An analogous analysis was also done examining the relationship between fTR and TETRAS scores of either tremor type before treatment, a proxy for their initial tremor severity (Supplementary Eq. S2 C,D, α=0.05).

### 1.4 Phase 2: Intra-operative Thalamic Recording with Concurrent TAPS Treatment

To assess if TAPS causally modulated broadband LFP or spiking activity in the contralateral VIM, we acquired thalamic microelectrode recordings during the mapping phase of each subject’s awake DBS surgery (Fig. 2D). Each subject avoided tremor medications for at least 24 hours before their operation. Prior to initial sedation, we also fitted a wrist stimulator on their dominant upper limb and verified median and radial hand dermatome paresthesia. Prior to the mapping phase, remifentanil sedation was discontinued for 15 minutes. During the mapping phase, neural activity from the hemisphere contralateral to the dominant upper limb (i.e. limb fitted with the wrist stimulator) was recorded from an array of 3 microelectrodes, 2 mm apart in the anterior-posterior axis, along a trajectory beginning 10 mm above the target. Here, the electrode in the center of the array was targeted to the VIM using consensus coordinates, 11 mm lateral to the wall of 3^rd^ ventricle at the level of the AC-PC plane, 5.7 mm posterior to the midcommissural point (i.e. x=12, y=-5.7, z=0; Fig. 3C).^60^ To map the thalamic nuclei, the borders of the ventralis oralis posterior, ventral intermediate, and ventral caudal nuclei were determined based on neural responses evoked by motor, kinesthetic, and somatosensory stimuli on the face and arms.^61^ At selected depths along trajectories planned for clinical mapping of the relevant nuclei, 60 seconds of microelectrode recordings (MER) was collected for each of two conditions: TAPS treatment ON and treatment OFF. With TAPS treatment ON, subjects were stimulated in for 60 seconds using their calibrated stimulation amplitude and alternating burst frequency from Phase 1. Bipolar neural recordings were made between each microelectrode in the array and separate low impedance macroelectrodes located 1 cm dorsal to the tip of the microelectrode. Signals were bandlimited at 20 kHz and digitized at 48 kHz using the Guideline 4000 LP+ (FHC, Inc, Bodwin, ME).

Following the mapping procedure, short bouts of electrical stimulation (biphasic, cathode leading 100 us pulses, 150 Hz, 0-4 mA current amplitude) were delivered through each of the 3 macroelectrodes to verify therapeutic efficacy of DBS and assess the resulting side effect profile. The final implant location for the DBS lead was chosen to be region where: 1) test stimulation resulted in the greatest tremor suppression, 2) test stimulation evoked minimal therapy limiting side effects (i.e. paresthesia, muscle contractions and dysarthria), and 3) MER revealed tremor cells and/or neural activity responsive to passive movement of the hand and arm. Putatively this describes the optimal placement of VIM DBS leads for the treatment of ET, projected to be in the ventral VIM 2-4 mm anterior to the border of the ventral caudal nucleus.^61^ For purposes of subsequent analyses, the final implant location is considered to be the coordinates that described the placement of the distal edge of the most distal contact on the DBS lead.

#### 1.4.2 Characterizing Thalamic Local Field Potentials

Prior to analysis, raw MER were low pass filtered at 7500 Hz and motion artifacts were removed with a custom algorithm that combined Banks et. al’s method to ours own (see Supplementary Methods 1.1, Supplementary Fig. S7).^62^ We also verified that turning TAPS ON did not induce stimulation artifact in thalamic MER (Supplementary Fig. S8) Then, each 60-second MER time series was segmented into 1-second bins and power spectral density (PSD) estimates were computed using Thomson’s multi-taper method with Slepian tapers (Matlab pmtm function, time-half bandwidth product = 3). Next, area under the curve (AuC) of the PSD, i.e. power, was calculated within canonical LFP frequency bands: delta (1-4 Hz), theta (4-8 Hz), alpha (8-12 Hz), beta (12-30 Hz), low gamma (30-70 Hz), and high gamma (70-200 Hz).^63–66^ For each LFP frequency band in each 60-second MER time series, power was taken as the median values after calculation was done for all 1-second bins. For each frequency band, we derived the LFP modulation index (MI_LFP_) to characterize how TAPS affect field potentials in the VIM. MI_LFP_ was quantified as a fractional change in power (PSD AuC) of the TAPS treatment ON versus OFF conditions (Fig. 3A, B):

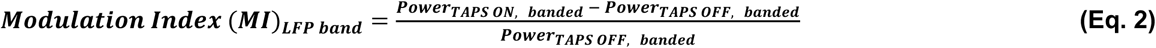

#### 1.4.3 Characterizing Thalamic Neural Spiking Activities

We also examined the effect of TAPS on spiking activity (see Supplementary Methods 1.2). Briefly, spike detection was performed by marking times where the preprocessed MER time series rises above 5 standard deviations from the series’ mean signal (Fig. 4A). Recordings with likely multiunit activity were further filtered for if their spike rate was at least 1 Hz during the treatment OFF condition. Next, spike times for each time series were binned (1 second bin width) and a custom bootstrap resampling procedure was used to estimate the distribution of spike rates (Fig. 4B). The firing rate of each example of multiunit activity was estimated as the median of the bootstrapped distribution. Bootstrap resampling was done to reduce bias from unequal time series duration after artifact removal. Similar to MI_LFP_, we calculated the modulation index for spiking activity (MI_Spikes_) to quantify fractional changes during the TAPS treatment ON versus OFF conditions (Fig. 4E, F):

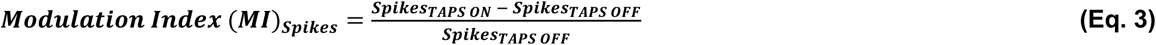

#### 1.4.4 Comparative Analysis

Linear regression models were used to examine the relationship between either MI_LFP_ or MI_Spikes_ and either the distance from the implant location (DIL, mm) (see Supplementary Methods 1.3) or fractional tremor reduction (fTR) (Supplementary Eq. S3, S4). Histologically, the VIM measures about 4 mm anterioposteriorly, 4 mm mediolaterally, and 6 mm dorsoventrally.^67–69^ Thus, we approximated the diameter of the VIM to be 5 mm and segregated MER data at this threshold. An α=0.05 error rate was used for MI_Spikes_ comparisons. For MI_LFP_ comparisons, we further applied a Bonferroni multi-comparison correction such that α’=0.008 was used as the most appropriate threshold of significance since regression for MI_LFP_ involved 6 bands. All analyses and statistical comparisons were performed in MATLAB 2024a (Mathworks, Natick, MA).

## Data availability

Raw neural data structures are not openly available due to the inclusion of private health information and are available from the corresponding author upon reasonable request. The de-identified data supporting the findings of this study will be shared on DRYAD once accepted for publication.

## Notes

### Competing Interest Statement

The authors have declared no competing interest.

### Clinical Trial

Study focused on testing for mechanism, not appropriate for clinical trial registration.

### Funding Statement

AJS and WBL received partial funding for this study from Cala Health, Inc.

### Author Declarations

The study was approved by the Institutional Review Board (IRB) at the University of Wisconsin, Madison (IRB ID: 2018-1052).

