## Supplemental Methods and Figures for "Transcutaneous afferent patterned stimulation reduces essential tremor symptoms through modulation of neural activity in the ventral intermediate nucleus of the thalamus"

*Supplementary Appendix*

1. Supplementary Methods
2. Supplementary Equations
3. Supplementary Figures

### 1. Supplementary Methods

#### *1.1 Motion Artifacts Removal*

Raw MER underwent zero-phase digital filtering (filtfilt function) using the transfer function coefficients of a 4th-order Butterworth filter, low-pass cutoff frequency at 7500 Hz.

Next, we applied a portion of Banks et. al's algorithm to create the first mask for blanking transient artifacts across all 3 recording channels at each depth.[1] "Transient artifacts were detected by identifying voltage deflections exceeding 10 standard deviations on a given channel. A time window was identified extending before and after the detected artifact until the voltage returned to the zero-mean baseline plus an additional 100 ms buffer before and after. High-frequency artifacts were also removed by masking segments of data with high gamma power exceeding 5 standard deviations of the mean across all segments." We reasoned that motion would induce artifacts in all channels, thus, we extended the algorithm to mark regions for blanking only when artifacts appeared across all 3 channels simultaneously.

While the algorithm could identify and blank portions of what we suspected as motion artifacts, on visual inspection, it left behind many obvious artifact segments between blanking regions. We created a second artifact removal mask using an iterative scanning window method to optimize for sensitivity and specificity. On a given channel, we calculated the moving root mean square (RMS) windowed at 0.5 ms. We tagged a point for removal if it exceeded 4 standard deviations of the mean moving RMS in the whole channel; this is the working masking. Next, we iteratively scanned in overlapping windows of 0.1, 25, and 250 ms and tagged all time point in the window for removal if it contained at least 2, 30, and 100 points, respectively, marked for removal. This iterative process increased sensitivity of artifact removal under the hypothesis that segments between RMS spikes that are close in time are likely motion artifact. We improved the specificity for motion artifact by creating a unified blanking mask that only blank segments where all 3 channels were tagged for removal. Additionally, we superimposed the 3 channels in the working mask to calculate the blanking density, a quotient dividing the number of points where RMS exceeded 4 standard deviations of mean RMS to their respective blanking window duration. We created the final second mask by filtering for blanking segments of at least 1 ms, with blanking density of at least 0.5 standard deviations above mean blanking density of all segments in the unified mask.

We merged the masks from Banks et al's algorithm and our iterative scanning window algorithm to create the third mask. However, we noticed that there were still gaps between blanking long blanking segments that were likely artifacts. We bridged them by identifying gaps that were 250-750 ms long and tagging them for blanking if the segment of the enveloping artifacts and gap has artifact that exceed 66% of the segment duration (Supplementary Fig. S8).

Prior to PSD or spike count calculation, we binned MER in consecutive 1 second segments. Segments with greater than 0.1 second of motion artifacts were further blanked and excluded from the analysis.

#### *1.2 Spikes Acquisition*

To extract spiking activities, raw MER underwent a similar filtering method as during motion artifact removal, though here we used a 4th order butterworth bandpass filter with cut-off frequencies between 500 and 7500 Hz. We removed time segment previously identified as motion artifact and identified spikes using a threshold-cross method. On a given channel, we normalized the signal by taking a difference of the signal and the mean across the recording duration. Spikes onsets were marked when the normalized signal exceeded 5 standard deviations of its mean. We removed consecutive spikes having inter-spike interval less than 1 ms as these are likely too frequent to be physiologic spikes.

Next, we filtered for areas with likely multiunit activity, selecting the recordings where spike rate was at least 1 Hz during the treatment OFF condition. Then, in each selected recording, we calculated the spike count in nonoverlapping 1 second segments. To increase the accuracy of our spike rate estimate, we applied a bootstrap resampling method that resampled the segments with replacement 10000 times to estimate the distribution of spike rates for each time series. The most likely spike rate was selected as the median from the distribution.

*1.3 Distance from Implant Location*

We defined the distance from implant location (DIL) as the Euclidean distance between each recording site and the final position of the bottom edge of the most inferior contact on the implanted electrode. This metric provides greater precision than depth measurements alone, which is crucial since the microelectrodes in our recording array are spaced 2 mm apart and likely detected varying amplitudes of Local Field Potentials (LFPs) and spike rates. While the initial DBS target in the ventral intermediate nucleus (VIM) is selected preoperatively using patient imaging data and standard stereotactic coordinates, the final implant location may differ based on intraoperative electrophysiological mapping and the observed balance between tremor reduction and side effects during stimulation testing. Therefore, characterizing neural activity relative to DIL, rather than the planned target, was more accurate site of actual treatment by DBS implant.

### 2. Supplementary Equations

$$\boldsymbol{Improvement_{total}\ (I)\ =\frac{1}{5}\sum_{k=1}^{5}(Task\ Score_{k,pre-treatment}-Task\ Score_{k,post-treatment})}$$ **(Eq. S1)**

**Equation S1.** Total tremor improvement was calculated by averaging changes across 5 TETRAS tasks (forward postural hold, lateral "wing beating" postural hold, kinetic finger-to-nose, spiral drawing, and dot approximation) in both treated and untreated limbs. Changes were calculated as pre-treatment minus post-treatment scores, where positive values indicate tremor improvement and negative values indicate tremor worsening.

$$\boldsymbol{fractional\ Tremor\ Reduction\ (fTR) \sim I_{kinetic} + I_{spiral\ drawing} + I_{dot\ approximation}}$$ **(Eq. S2A)**

$$\boldsymbol{fractional\ Tremor\ Reduction\ (fTR) \sim I_{forward} + I_{lateral}}$$ **(Eq. S2B)**

$$\boldsymbol{fractional\ Tremor\ Reduction\ (fTR) \sim S_{kinetic} + S_{spiral\ drawing} + S_{dot\ approximation}}$$ **(Eq. S2C)**

$$\boldsymbol{fractional\ Tremor\ Reduction\ (fTR) \sim S_{forward} + S_{lateral}}$$ **(Eq. S2D)**

**Equation S2.** Multiple linear regression models examining relationships between fTR and: **(S2A)** improvements in kinetic tremors, **(S2B)** improvements in postural tremors, **(S2C)** baseline kinetic tremor TETRAS scores, and **(S2D)** baseline postural tremor TETRAS scores.

$$\boldsymbol{Modulation\ Index\ (MI)_{LFP-Band} \sim Distance\ from\ Implant\ Location\ (DIL)}$$ **(S3A)**

$$\boldsymbol{Modulation\ Index\ (MI)_{LFP-Band} \sim functional\ Tremor\ Reduction\ (\mathrm{fTR})}$$ **(S3B)**

**Equation S3.** Simple linear regression models studying how TAPS-induced modulation of local field potentials across canonical frequency bands relates to **(S3A)** DIL and **(S3B)** fTR.

$$\boldsymbol{Modulation\ Index\ (MI)_{Spikes} \sim Distance\ from\ Implant\ Location\ (DIL)}$$ **(S4A)**

$$\boldsymbol{Modulation\ Index\ (MI)_{Spikes} \sim functional\ Tremor\ Reduction\ (\mathrm{fTR})}$$ **(S4B)**

**Equation S4.** Simple linear regression models studying how TAPS-induced modulation of spiking activity relates to **(S4A)** DIL and **(S4B)** fTR.

**3. Supplementary Figures**

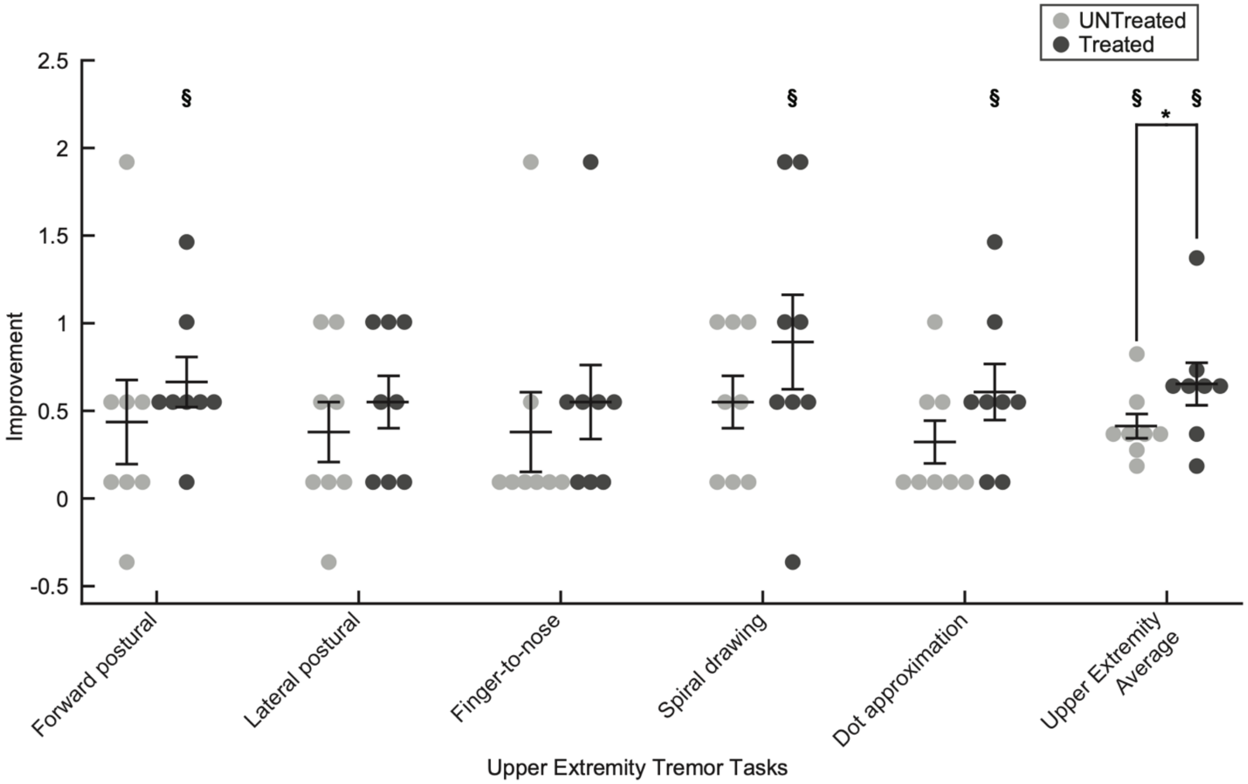


**Figure S1**. **TAPS improved tremor in both limbs, with only the dominant, treated limb showing significant improvements in specific tasks.** In the treated limb, forward postural hold tremor improved by 0.625 ± 0.157 ($^{§}p=0.016$), spiral drawing by 0.875 ± 0.295 ($^{§}p=0.039$), and dot approximation by 0.563 ± 0.175 ($^{§}p=0.031$). Average total tremor improvement was 0.613 ± 0.133 ($^{§}p=0.008$) in the treated limb and 0.35 ± 0.076 ($^{§}p=0.008$) in the untreated limb, with a significant difference between limbs (*p=0.047). All values show Mean ± SEM. $^{§}$One-sample, two-sided, Wilcoxon signed-rank test with p≤0.05. *Two-sample, two-sided, Wilcoxon signed-rank test with p≤0.05.

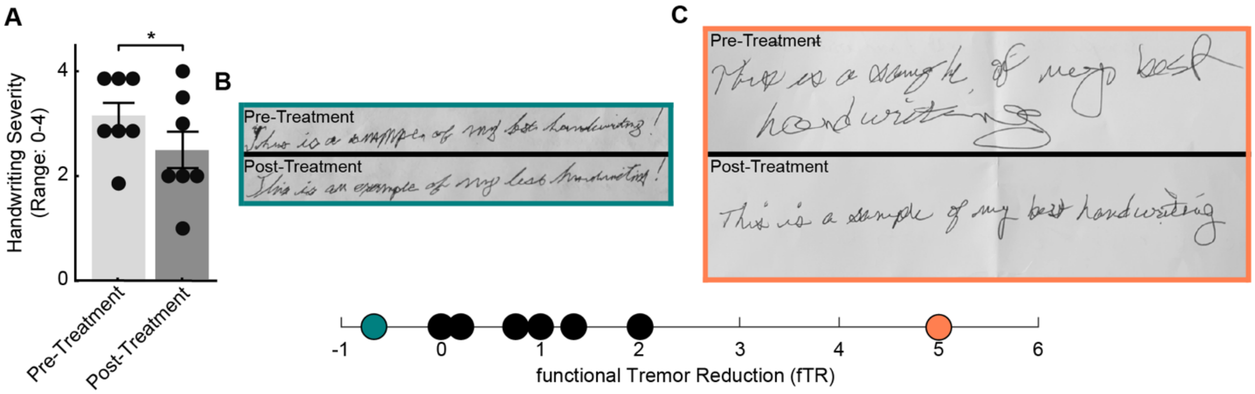


**Figure S2. Handwriting tremor in the dominant limb significantly improves following TAPS treatment (n=7). A** Handwriting from the dominant, treated limb showed pre-treatment scores (Mean ± SEM = 3.29 ± 0.29) decreased relative to post-treatment scores (2.50 ± 0.39) (*two-sided signed-rank test with p=0.002). Data was not collected one patient who writes with their non-dominant, untreated limb. **B–C** Representative handwriting samples before and after TAPS treatment from patients with **(B)** low (cyan) and **(C)** high (orange) fTR.

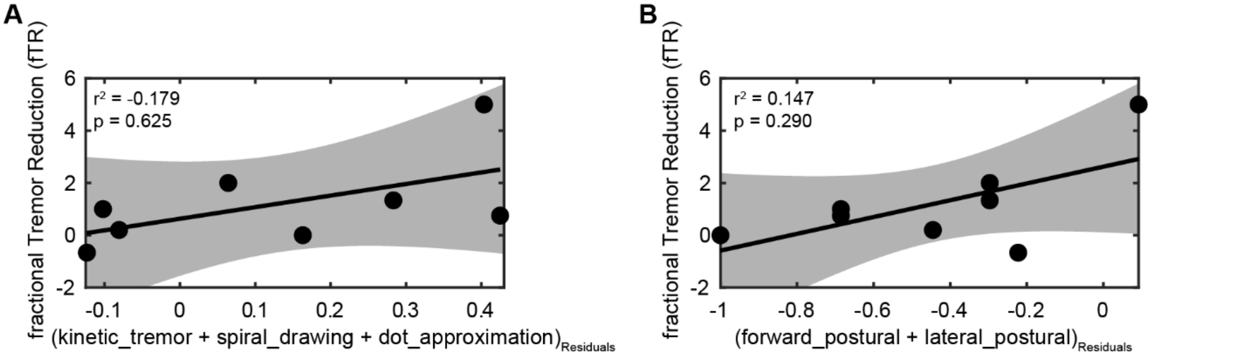


**Figure S3. Initial kinetic and postural tremor scores show no significant correlation with fTR. A–B** Multiple linear regression analyses showing the relationship between fTR and baseline TETRAS scores for **(A)** kinetic tremor tasks and **(B)** postural tremor tasks.

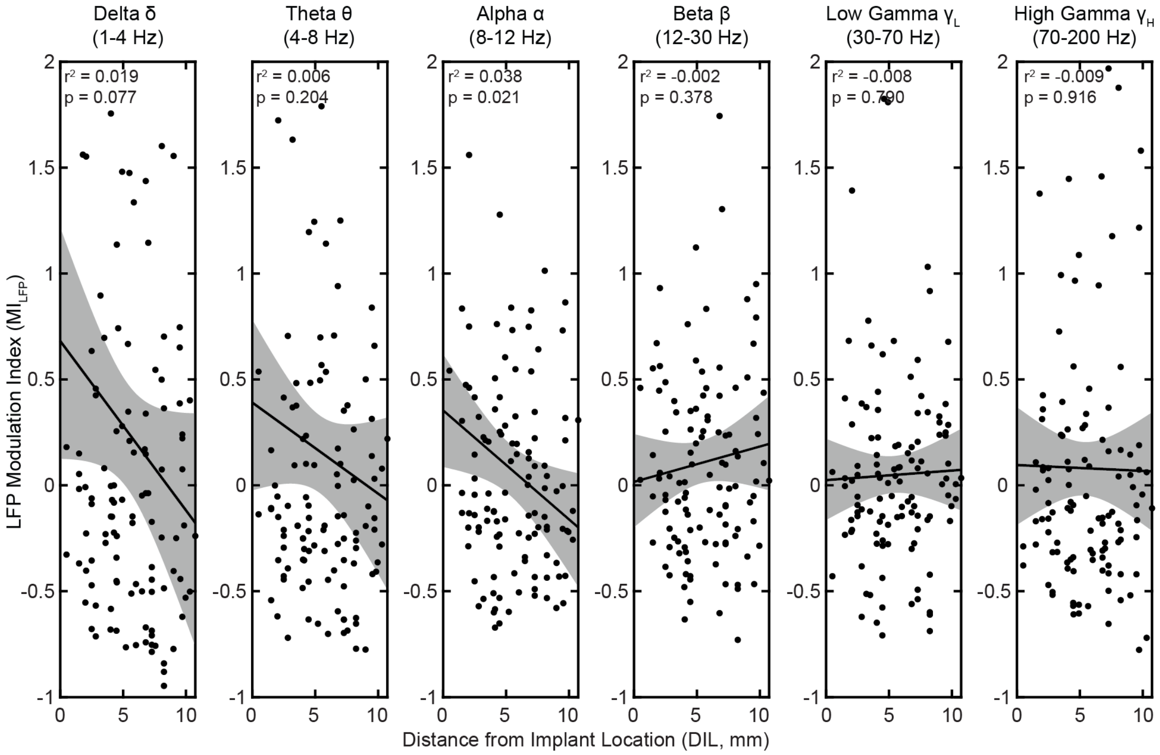


**Figure S4. LFP modulation ($MI_{LFP}$) shows no correlation with distance from implant location (DIL) when data from 0-10 mm is considered together.** Linear regression analyses comparing $MI_{LFP}$ to DIL across 6 frequency bands. Unlike Supplementary Fig. S5, MER data was analyzed together without segregation based on location that is within or outside the VIM. The α-band modulation showed a trend toward significance but did not meet the Bonferroni-corrected threshold (α'=0.008).

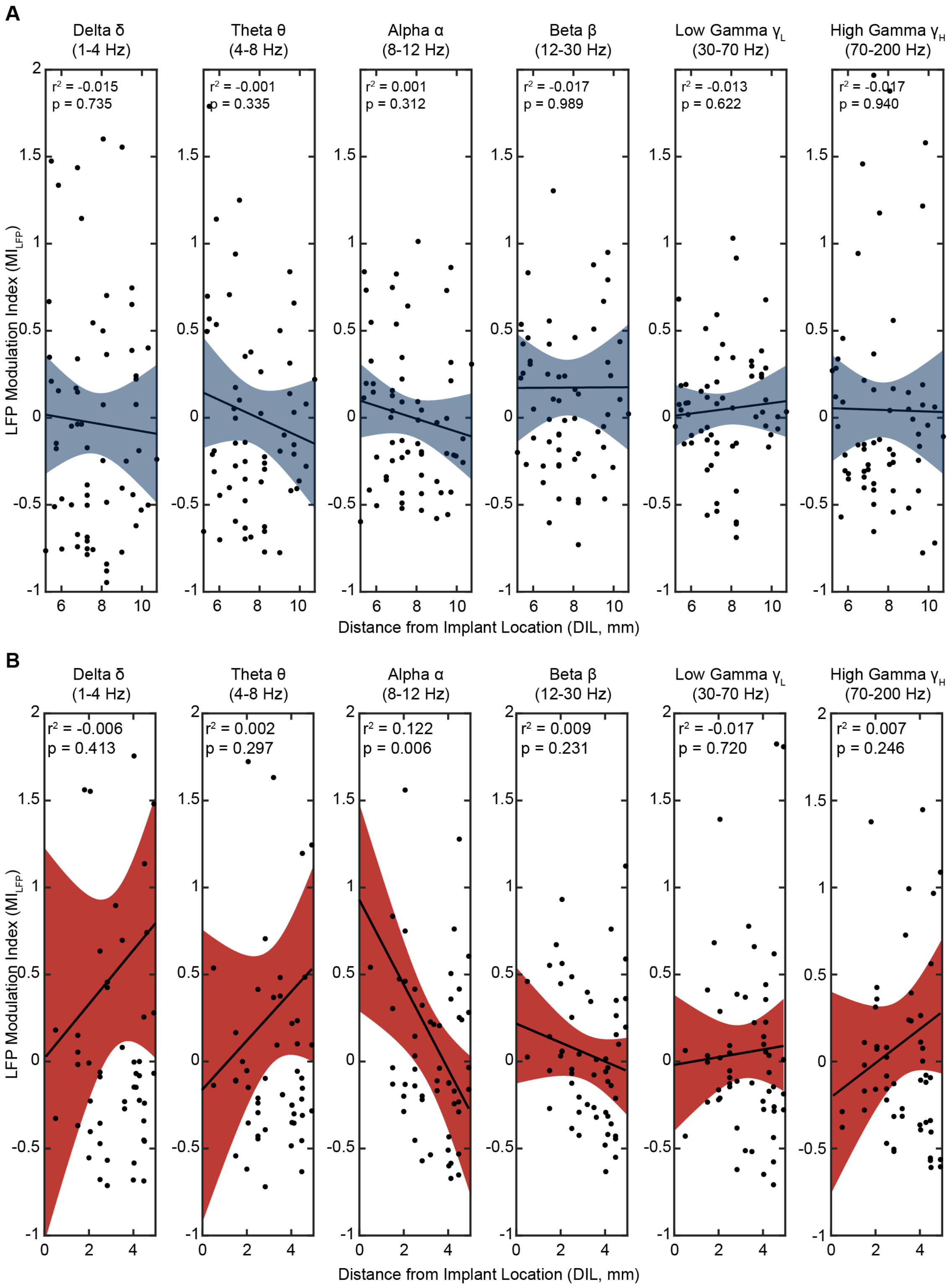


**Figure S5. Within the VIM, $MI_{LFP}$ displays a significant, negative correlation to DIL for the alpha band. A–B** Linear regression analyses comparing $MI_{LFP}$ to DIL across 6 frequency bands for recordings **(A)** outside VIM (5.01-10 mm, blue) and **(B)** within VIM (0-5 mm, red). Significance threshold was Bonferroni-corrected to α'=0.008.

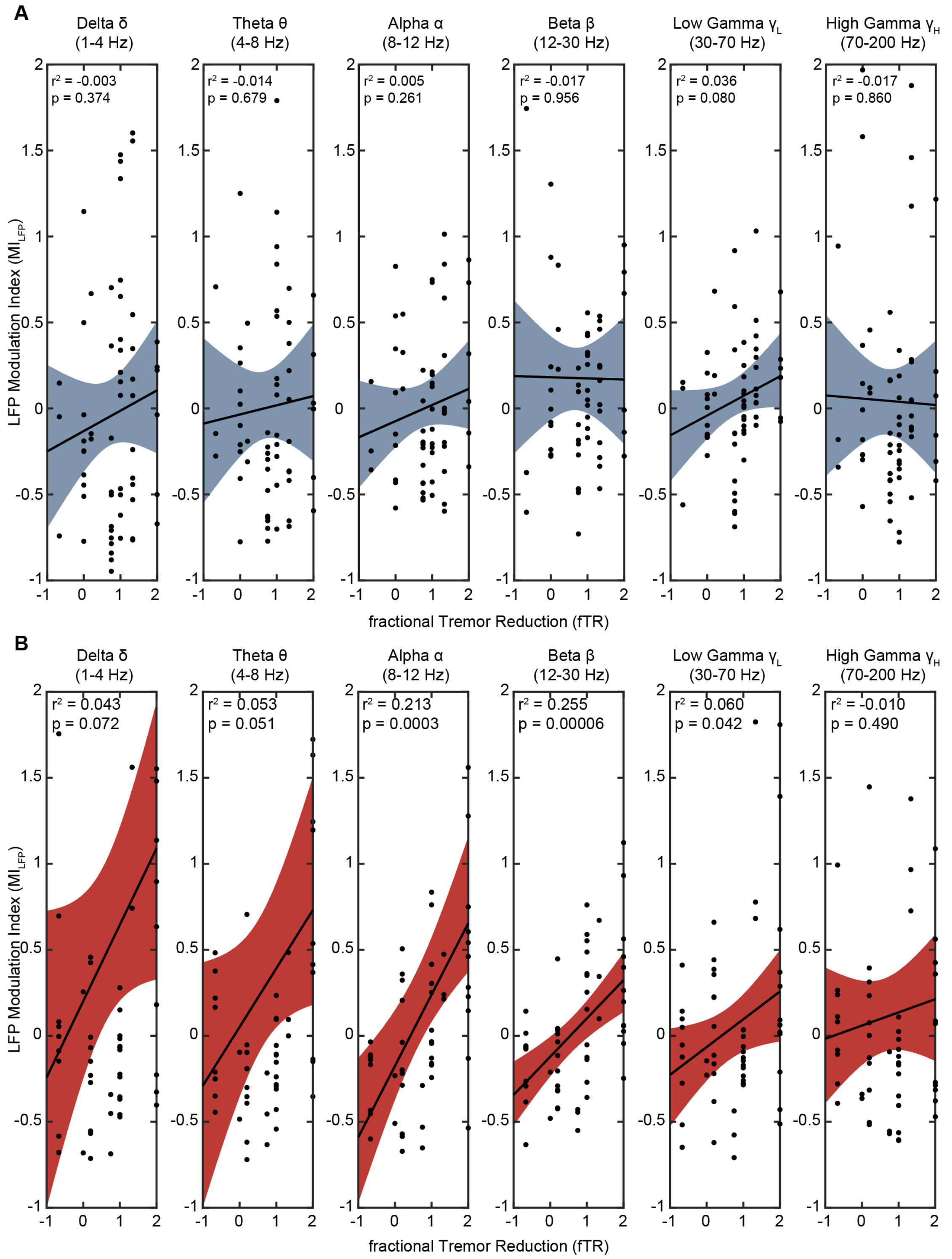


**Figure S6. Within the VIM, $MI_{LFP}$ displays a significant, positive correlation to fTR for the alpha and beta bands. A–B** Linear regression analyses comparing $MI_{LFP}$ to fTR across 6 frequency bands for recordings **(A)** outside VIM (5.01-10 mm, blue) and **(B)** within VIM (0-5 mm, red). Significance threshold was Bonferroni-corrected to α'=0.008.

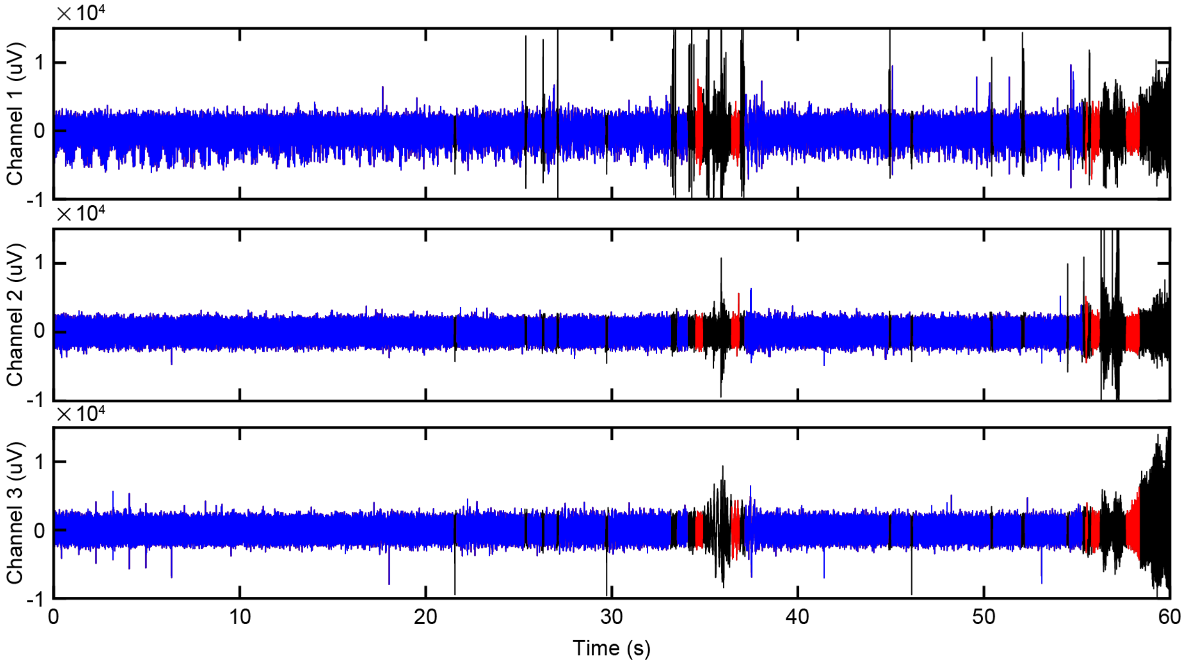


**Figure S7. Motion artifacts from thalamic MER were removed prior to further analysis.** Sample microelectrode recordings (MER) taken 7.50-7.76 mm from the final implant location, showing recordings from the anterior (Channel 1), middle (Channel 2), and posterior (Channel 3) electrodes. Low-pass filtered waveforms (blue) are shown with artifact segments (black) identified using combined criteria from Bank et al. and our iterative scanning window masks. Signal segments between artifacts were further marked for blanking (red) if artifacts make up at least 66% of the segment created by including adjacent artifacts.

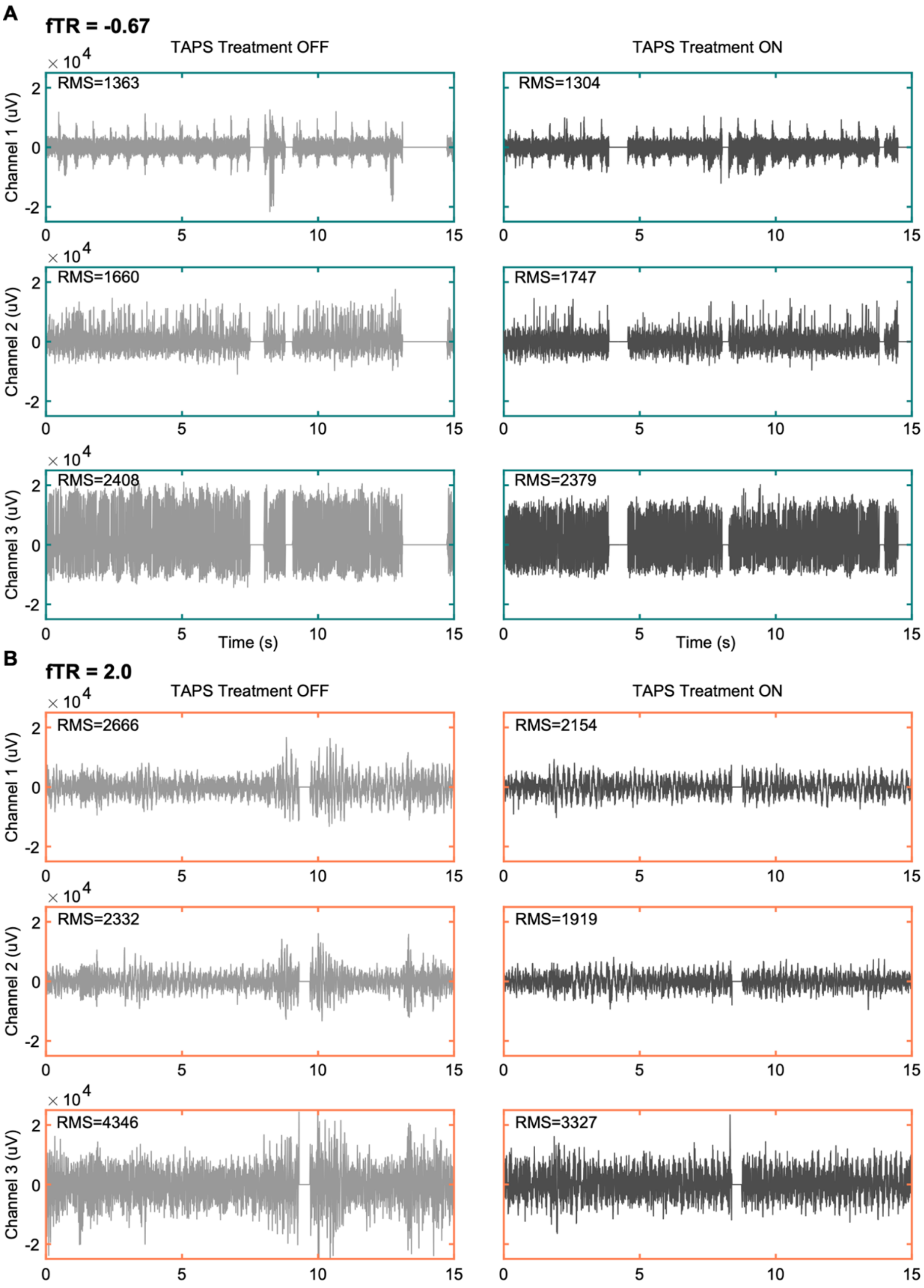


**Figure S8. TAPS did not produce stimulation artifacts in MER data, regardless of low or high fTR. A–B** Representative multi-channel recordings taken 3 mm from target in two patients with different treatment responses: **(A)** low fTR (-0.067) and **(B)** high fTR (2.0). Fifteen-second segments from 60-second recordings are shown to enable detailed visual inspection. Stimulation artifacts, not seen, would appear as an additive pattern of regular spikes during the TAPS ON state, increasing the RMS value. Segments of no signal (0 uV) concurring across 3 channels are segments of motion artifacts, removed by Supplementary Methods 1.1.
